# Artificial Intelligence in Colposcopy: Evaluation of ChatGPT as a Diagnostic and Predictive Support Tool

**DOI:** 10.1101/2025.04.19.25326103

**Authors:** Carlos Piñel Pérez, María José Gómez-Roso Jareño

## Abstract

**Background:** Colposcopy remains a critical step in the diagnostic algorithm following a positive HPV test, but it suffers from limitations such as inter-operator variability and reliance on clinical expertise. The integration of artificial intelligence (AI) could enhance diagnostic consistency and support decision-making, especially in resource-limited settings.

**Objective:** To evaluate the performance of ChatGPT, a generative AI language model, in the diagnostic interpretation of colposcopic images, and to compare its clinical decisions and histological predictions with those of expert colposcopists and real pathology outcomes.

**Methods:** A retrospective study was conducted using 146 colposcopic cases sourced from the publicly available IARC Colposcopy Atlas. For each case, ChatGPT was provided with static images, patient age, and HPV status and asked to classify the case (physiological vs. pathological), recommend a clinical action (biopsy or not), and predict the histological outcome. The results were compared with expert colposcopist decisions and pathology reports.

**Results:** There was a high agreement between ChatGPT and colposcopists in diagnostic impression (92.5%, κ = 0.83) and biopsy decision (82.9%, κ = 0.58). In biopsied cases (n = 95), ChatGPT matched the histological diagnosis in 88.4% of cases. Sensitivity and positive predictive value for detecting CIN2+ were 97.1% and 94.4%, respectively. In logistic regression models predicting CIN2+, ChatGPT-based models achieved comparable AUCs to colposcopist-based models (0.947 vs. 0.966, p = 0.129).

**Conclusion:** ChatGPT demonstrated high concordance with expert evaluations and robust diagnostic performance. Its multimodal reasoning capabilities and accessibility suggest that language-based AI models may serve as valuable support tools in colposcopy, particularly where expertise is limited.

## INTRODUCTION

The World Health Organization (WHO) aims to eliminate cervical cancer as a public health problem through the universal implementation of effective vaccination, screening, and treatment strategies. Routine cervical cancer screening significantly reduces mortality by enabling early detection of precancerous lesions and allowing timely treatment (2,3,4). HPV testing is more sensitive than cytology in detecting cervical precancer and provides greater long-term reassurance when the result is negative (5,6). Colposcopy is the reference standard for diagnosing cervical precancer and cancer in HPV-positive women, guiding biopsy and treatment decisions (6). It is used to evaluate women with positive HPV tests, ensuring that high-risk cases are appropriately identified and managed (7,8).

However, colposcopy presents several limitations that affect its effectiveness. It relies heavily on the subjective experience of the operator, leading to significant inter- and intra-observer variability. This dependency can result in inconsistent interpretations and reporting, especially in settings where experienced colposcopists are scarce (9,10). The skill required for effective colposcopy cannot be easily acquired or maintained without regular clinical practice in high-volume settings with abnormal cases. This is challenging due to the low incidence of invasive disease in the general population, which can lead to diagnostic errors, particularly in detecting microinvasion (11). The sensitivity of colposcopy to detect high-grade cervical intraepithelial neoplasia has been reported as low as 54% to 57% in some studies. This is partly due to the difficulty in identifying small high-grade lesions, which are increasingly detected through more sensitive screening methods (12).

Artificial intelligence (AI) plays a significant role in medical image analysis and clinical decision-making by enhancing diagnostic accuracy, efficiency, and decision support across various medical fields. AI, particularly deep learning algorithms such as convolutional neural networks, excels at recognizing complex patterns in medical images, providing quantitative assessments that surpass traditional qualitative evaluations by physicians (13,14,15). AI systems, when combined with clinician expertise, enhance diagnostic confidence and overall system performance, although they do not achieve 100% accuracy (16). Moreover, AI supports clinical decision-making by optimizing care pathways for chronic diseases, suggesting precision therapies, and improving patient management (16,17).

Within this context, there is growing interest in exploring whether advanced and accessible AI models such as ChatGPT can complement—or even match—the diagnostic decision-making of expert colposcopists. Given its potential to interpret clinical images and reason contextually, its application in colposcopic analysis represents a promising opportunity, particularly in settings with limited specialized expertise or high diagnostic variability.

The aim of this study was to evaluate the performance of ChatGPT in analyzing colposcopic images by comparing its diagnostic impression, clinical attitude (biopsy decision), and histological prediction to those of expert colposcopists, as well as to actual histopathological results when available.

## MATERIALS AND METHODS

### Study design

A retrospective observational study was conducted to evaluate the concordance and diagnostic performance of a natural language-based artificial intelligence model (ChatGPT, OpenAI) in the analysis of colposcopic images. The primary objective was to compare the model’s performance with that of expert colposcopists and with available histopathological results. The study was carried out between January and April 2025.

### Data source

The colposcopic images were obtained from the public and freely accessible repository *Atlas of Colposcopy – Principles and Practice*, developed by the International Agency for Research on Cancer (IARC), under the auspices of the World Health Organization (WHO) (18).

This resource includes high-quality colposcopic images accompanied by basic clinical data (age, HPV status), expert colposcopist impressions, and histopathological outcomes for biopsied cases. The database is intended for clinical training and allows unrestricted access for academic and research purposes.

### Case selection

A total of 146 unique cases with diagnostically adequate colposcopic images were consecutively included, manually selected based on the following inclusion criteria: images classified under the “Normal,” “Metaplasia,” “Low-grade,” “High-grade,” or “Cancer” sections; availability of acetic acid-applied colposcopic images (and Lugol-stained images when available); associated clinical data including age and HPV test result; expert colposcopist’s diagnostic impression (physiologic or pathologic); clinical decision on whether to perform a biopsy; and availability of histopathological result if a biopsy was performed.

Duplicate images, cases with insufficient clinical data, and low-quality images that precluded reasonable interpretation were excluded.

### Evaluation by the AI model (GPT)

ChatGPT (GPT-4 model by OpenAI, April 2024 release) was instructed to analyze the images blindly, without access to original interpretations or histological results. For each case, the following information was provided: the acetic acid-stained colposcopic image (and Lugol-stained if available), the patient’s age, and HPV status. Cytology results, histological findings, and expert impressions were not disclosed. For each case, ChatGPT was asked to provide: a visual diagnostic impression (non-significant vs. pathologic lesion), a clinical recommendation (biopsy vs. no biopsy), a predicted histological diagnosis (if a biopsy were to be performed). All responses were recorded in a structured database, and no feedback or cross-case learning was permitted.

### Variables and coding

For each case, the following clinical variables were recorded: patient age (continuous variable) and HPV test result (classified as positive or negative). Diagnostic impressions from both the expert colposcopist and ChatGPT were categorized as either non-significant (normal or metaplastic findings) or pathologic (suggestive of lesion).

Regarding clinical management, it was documented whether both the colposcopist and GPT recommended performing a biopsy based on the colposcopic image. These decisions were classified as “biopsy” or “no biopsy.” For cases in which a biopsy was performed, the definitive histological diagnosis was collected and grouped into five categories: normal, metaplasia, low-grade cervical intraepithelial neoplasia (CIN1), high-grade CIN (CIN2–3), and invasive carcinoma.

For concordance and diagnostic performance analyses, a binary variable was created—”significant lesion” (CIN2+)—which included all high-grade lesions (CIN2, CIN3, and cancer). This cutoff was chosen as it represents the usual clinical threshold for treatment. Cases without biopsy were specifically identified and excluded from analyses requiring histological confirmation.

### Statistical analysis

Statistical analyses were performed using Python 3.10 (with the libraries pandas, statsmodels, sklearn, and matplotlib), with parallel validation in Jamovi 2.4. Descriptive statistics of the sample were obtained. Diagnostic concordance between ChatGPT and expert colposcopists was evaluated using Cohen’s kappa coefficient for both diagnostic impression (pathologic vs. non-significant) and biopsy decision. A Venn diagram was generated to illustrate the overlap between biopsy decisions and histologically confirmed lesions.

A direct comparison between the histological diagnosis predicted by ChatGPT and the actual pathology report was performed by calculating the exact match rate. Diagnostic performance metrics for ChatGPT—including sensitivity, specificity, positive predictive value (PPV), and negative predictive value (NPV)—were computed for CIN1 and CIN2+ as diagnostic thresholds. A radial performance chart was also generated.

Two logistic regression models and receiver operating characteristic (ROC) curves were constructed to predict CIN2+ lesions: a clinical model (Age + HPV + colposcopist impression) and a GPT-based model (Age + HPV + GPT impression). Both models were applied to the same patient cohort, allowing for direct comparison of their area under the curve (AUC) using a two-tailed Z-test with bootstrapping.

## RESULTS

A total of 146 cases of patients undergoing colposcopy were analyzed, with ages ranging from 25 to 60 years (mean: 42.1 years; SD: 8.83). HPV infection was positive in 65.75% of patients. Expert colposcopists classified the colposcopy as pathological in 63.7% of cases, whereas ChatGPT considered 69.86% of cases as pathological. Colposcopists decided to biopsy in 65.07% of cases, compared to 80.82% of cases in which GPT recommended biopsy. Table 1 shows the histopathological results of the biopsied cases.

**Table 1.** Histopathological Diagnosis of Analyzed Cases.

| Histopathological Diagnosis | Frequency | Percentage |
| --- | --- | --- |
| No biopsy performed | 51 | 34.93% |
| Normal | 3 | 2.05% |
| Metaplasia | 6 | 4.11% |
| CIN 1 | 17 | 11.64% |
| CIN 2–3 | 45 | 30.82% |
| Cancer | 24 | 16.44% |

There was agreement between colposcopists and ChatGPT regarding the diagnostic impression in 135 out of 146 cases (raw agreement rate: 92.47%; 95% CI: 87.01%–95.74%), and in 121 out of 146 cases regarding the decision to biopsy or not (raw agreement rate: 82.88%; 95% CI: 75.94%–88.12%). Concordance between diagnostic impressions (pathological vs. non-significant findings) was very high, with a Cohen’s kappa of 0.83 (95% CI: 0.74–0.93). Concordance in biopsy decisions showed moderate agreement, with a kappa of 0.58 (95% CI: 0.44–0.72).

Among the 95 biopsied cases, ChatGPT correctly predicted the histological diagnosis in 84, achieving an exact match rate of 88.42% (95% CI: 80.45%–93.41%). Table 2 and Figure 2 present the diagnostic performance of the GPT model in these biopsied cases. CIN1 and CIN2+ (including CIN2–3 and cancer) were evaluated as separate diagnostic thresholds.

**Table 2.**
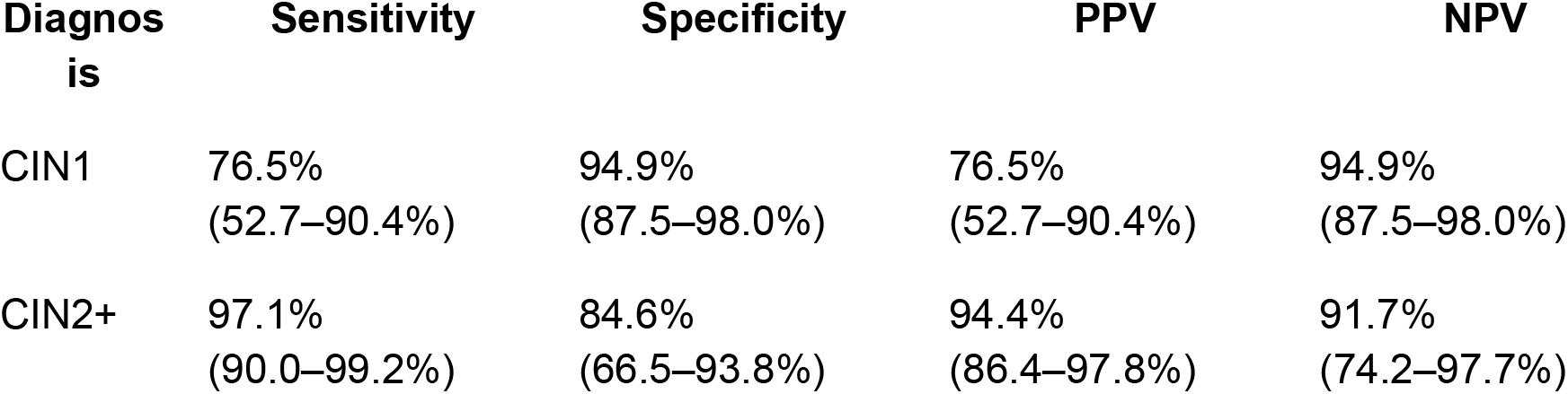
Diagnostic Performance of ChatGPT by Clinical Category (Biopsied Cases, N=95)

| Diagnosis | Sensitivity | Specificity | PPV | NPV |
| --- | --- | --- | --- | --- |
| CIN1 | 76.5%<br>(52.7–90.4%) | 94.9%<br>(87.5–98.0%) | 76.5%<br>(52.7–90.4%) | 94.9%<br>(87.5–98.0%) |
| CIN2+ | 97.1%<br>(90.0–99.2%) | 84.6%<br>(66.5–93.8%) | 94.4%<br>(86.4–97.8%) | 91.7%<br>(74.2–97.7%) |

**Figure 1.**
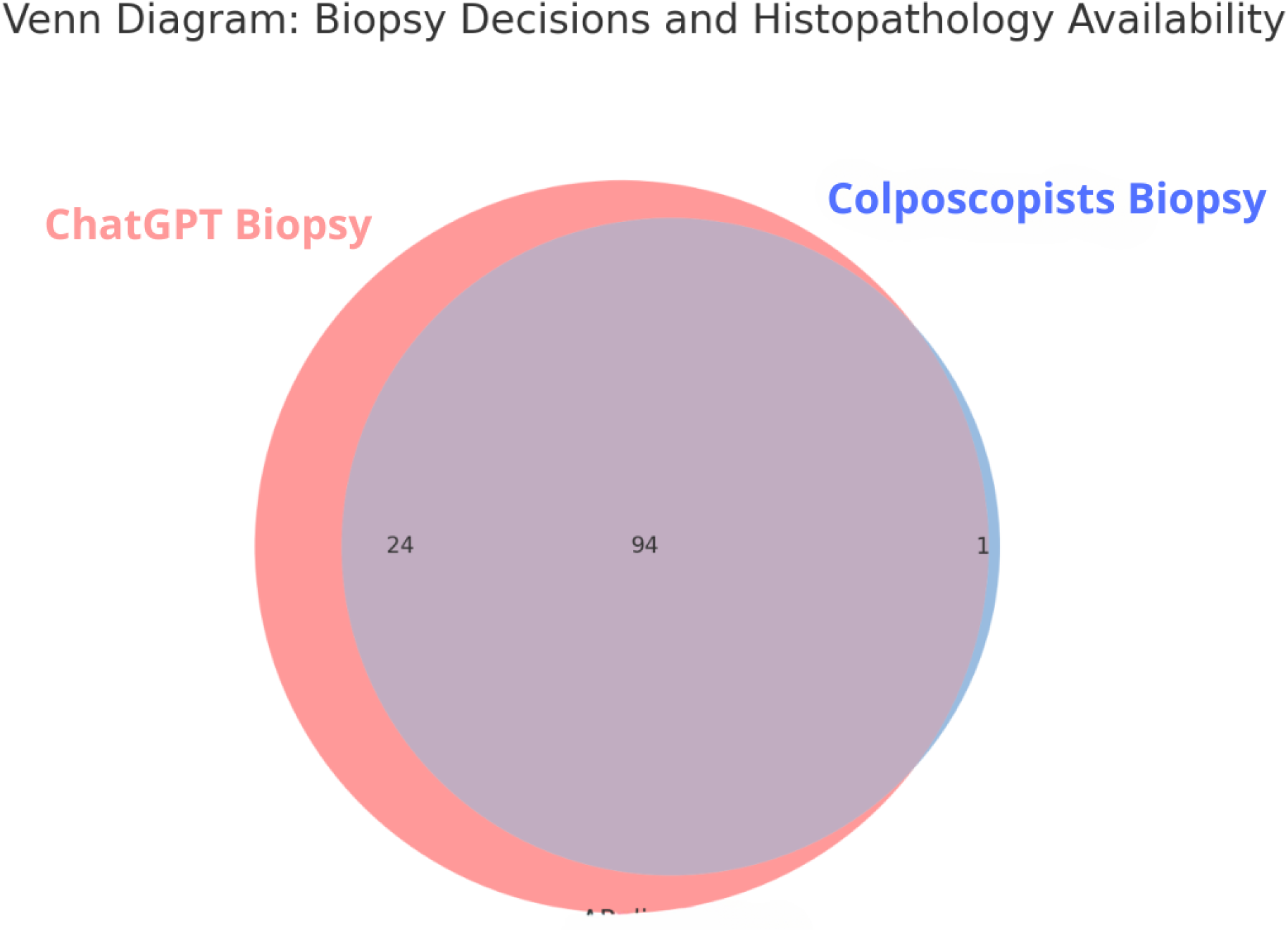
Venn diagram illustrating the intersection between the biopsy decisions of colposcopists (blue), those recommended by GPT (red). The central area (94 cases) represents agreement between GPT and the colposcopists in recommending biopsy and obtaining a pathology result. The 24 cases in the red sector indicate additional biopsies that GPT would have recommended but were not performed by the colposcopists. The single case in the blue area represents a biopsy performed by the colposcopist that GPT would not have indicated.

**Figure 2.**
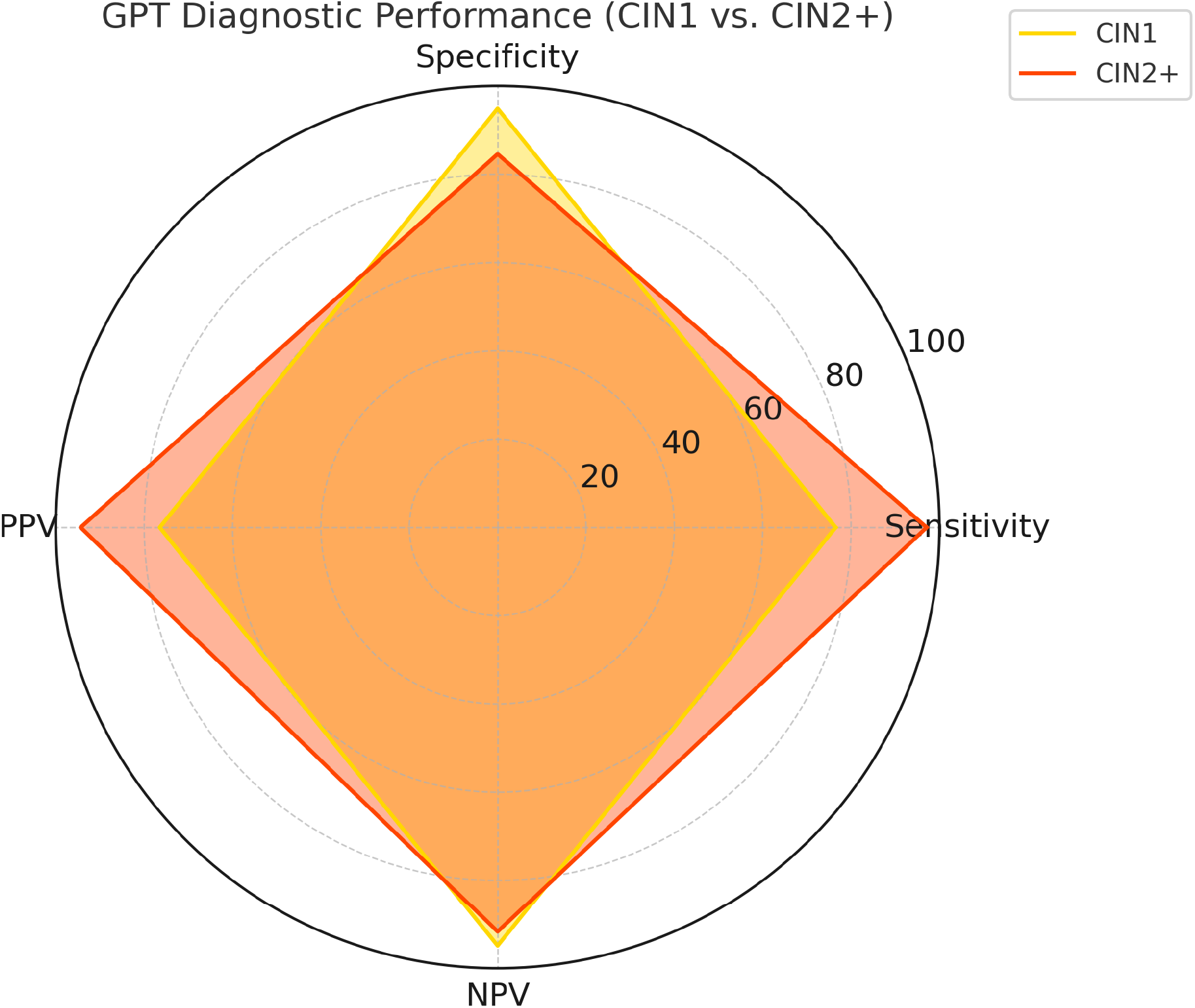
Radar chart comparing the diagnostic performance of the GPT model in detecting low-grade lesions (CIN1) and high-grade lesions (CIN2+). Sensitivity, specificity, positive predictive value (PPV), and negative predictive value (NPV) are shown for each diagnostic category, calculated from biopsied cases. The model demonstrated excellent performance in detecting clinically significant lesions (CIN2+), with a distinct performance profile for CIN1, characterized by high specificity and NPV.

Two logistic regression models were constructed to predict the presence of clinically significant cervical lesions (CIN2+). The clinical model included patient age, HPV status, and the colposcopist’s diagnostic impression as independent variables. The comparative model included the same variables, replacing the colposcopist’s impression with that of ChatGPT. Both models demonstrated excellent performance. The clinical model achieved an area under the curve (AUC) of 0.966 (95% CI: 0.934–0.990), while the GPT-based model yielded an AUC of 0.947 (95% CI: 0.908–0.977). The difference between the AUCs was not statistically significant (ΔAUC = 0.019; 95% CI: –0.006–0.045; p = 0.129). The corresponding ROC curves are shown in Figure 3.

**Figure 3.**
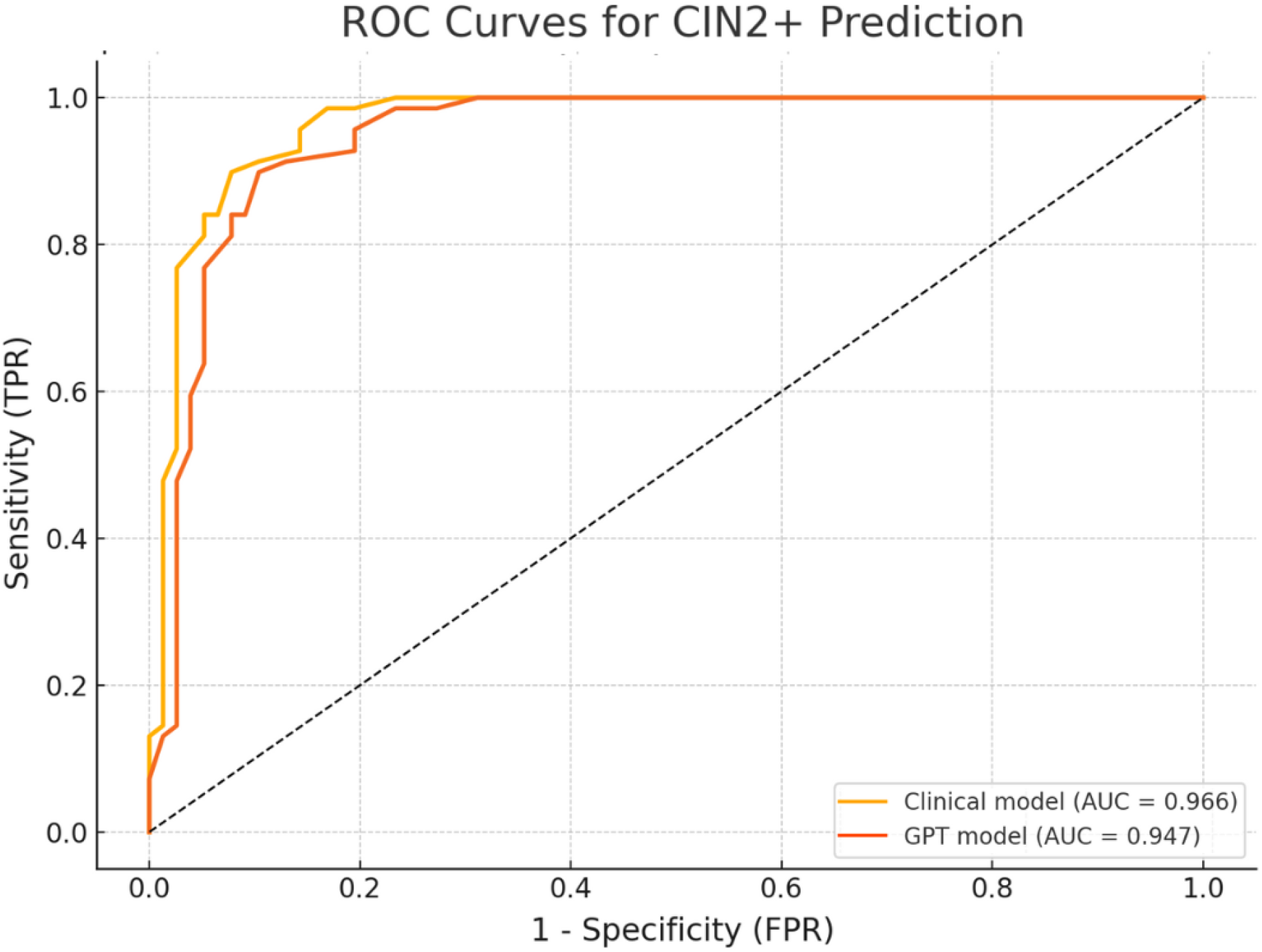
ROC curves of the logistic regression models for predicting high-grade lesions (CIN2+) based on clinical variables. The clinical model included age, HPV status, and the diagnostic impression of the colposcopist, while the comparative model replaced the colposcopist with GPT’s evaluation. Both models demonstrated excellent predictive capacity, with AUCs of 0.966 and 0.947, respectively.

## DISCUSSION

This study evaluated the diagnostic performance of a generative language model (ChatGPT) for colposcopic analysis, comparing it with expert colposcopists and using histopathology as the reference standard. A high level of agreement was observed between ChatGPT and expert colposcopists in the visual interpretation of colposcopic images. In over 92% of cases, ChatGPT matched the clinical judgment of the specialist in classifying the cervix as either pathological or without relevant lesions, with a very high concordance (Cohen’s kappa = 0.83).

Similarly, substantial agreement was found in the clinical decision to biopsy, with a concordance rate of 82.88%. In this case, the kappa coefficient was moderate (k = 0.58), since ChatGPT recommended biopsy in some cases it considered likely to be non-significant (e.g., metaplasia), whereas the colposcopists opted not to biopsy (Figure 1). This discrepancy may reflect a difference in decision-making approach: while the expert relies on experienced clinical judgment, ChatGPT may prefer to confirm the absence of premalignant lesions, considering that visual differences between metaplasia and low-grade lesions can sometimes be subtle and difficult to distinguish.

Among the 95 biopsied cases, ChatGPT correctly predicted the histological diagnosis in 88.4% of cases. Its performance for detecting high-grade lesions (CIN2+) was particularly impressive, achieving a sensitivity of 97.1% and a positive predictive value (PPV) of 94.4%. These results support its potential to accurately identify clinically significant lesions.

Furthermore, multivariable logistic regression models showed that ChatGPT, when integrated as a diagnostic evaluator, achieved predictive capability comparable to that of models based on colposcopist judgment. The difference between the areas under the ROC curve (AUC = 0.947 vs. 0.966) was not statistically significant, suggesting that ChatGPT could provide equivalent clinical support in diagnostic settings.

This multimodal reasoning approach and transparent interpretability may help explain the high diagnostic alignment between ChatGPT and human experts. Its ability to approximate histological outcomes further strengthens its role as a potential adjunctive tool, especially in settings with limited colposcopic expertise.

In this context, ChatGPT could become a valuable resource for trainees or environments with limited access to experienced colposcopists. Its use as an automated second-opinion system may help reduce diagnostic variability, standardize decision-making, and ultimately improve screening quality.

Likewise, in resource-limited regions—where access to high-quality screening programs or specialists may be constrained—an accessible AI tool like ChatGPT could help promote equity in diagnostic access, reducing gaps in early detection of cervical cancer.

The findings of this study align with recent research on AI in colposcopy, though with some notable distinctions. Xue et al. (2020) reported that the CAIADS system achieved 82.2% agreement with histology compared to 65.9% for colposcopists (kappa = 0.75 vs. 0.52) (19). Similarly, Medina et al. (2023) found that AI-assisted video colposcopy achieved 80% overall accuracy compared to 65% for conventional colposcopy (p < 0.001) (20). In our analysis, ChatGPT achieved a diagnostic concordance of 88.4% in biopsied lesions—slightly higher than published results for other AI systems and above the average performance of traditional colposcopy. However, our retrospective study was based on selected cases with didactic value, which may overestimate ChatGPT’s predictive power. Its real-world performance may be lower in more ambiguous or less illustrative cases.

Another relevant aspect is the sensitivity for detecting high-grade lesions (CIN2+). Our results indicate that ChatGPT identified 97.1% of confirmed CIN2+ lesions, meeting the clinical objective of avoiding missed precancerous lesions. The literature consistently shows that AI-based vision algorithms increase sensitivity for HSIL/CIN2+ compared to standard colposcopy. For example, CAIADS reached 90–91% sensitivity (depending on the diagnostic threshold used), compared to 83% among experts (19). Similarly, Medina et al. reported 96% sensitivity with AI-assisted video colposcopy versus 93% with conventional evaluation (20).

Regarding specificity, our study observed some differences. ChatGPT classified more cases as pathological compared to colposcopists (69.9% vs. 63.7%), suggesting a tendency to overdiagnose minor abnormalities to avoid missing serious lesions. This led to a higher proportion of biopsies recommended by ChatGPT (80.82%) compared to human experts (65.07%), increasing the number of potentially unnecessary biopsies. Consequently, specificity for CIN2+ was slightly lower with ChatGPT (84.6%) than reported in prior studies for expert performance, which ranges between 87.7% and 92.0% (21,22). This reflects the classic tradeoff between sensitivity and specificity: like other AI systems, ChatGPT prioritizes sensitivity at the potential cost of more false positives. Still, combining AI interpretations with human colposcopists has been shown to improve sensitivity while maintaining comparable specificity (23).

Deep learning models have been developed to classify cervical neoplasms from colposcopic images, demonstrating high accuracy in identifying lesions that require biopsy (24,25,26). When comparing ChatGPT’s performance with systems like CAIADS (19,27) or other deep learning models, we observed similar diagnostic metrics but substantial methodological differences.

A key difference lies in the analytical approach. While classical visual AI models process image pixels directly to segment lesions or suggest biopsy sites (25), ChatGPT reasons based on visual patterns and clinical context (e.g., patient age, HPV status) to generate a holistic diagnostic impression. It acts more like a virtual clinical expert than a traditional image-processing algorithm. This ability to integrate multiple data sources aligns with current clinical guidelines (28,29). Moreover, ChatGPT’s use of natural language enables explainable diagnostics—something traditional black-box AI models cannot offer.

Nevertheless, ChatGPT also has limitations. Unlike dedicated image-based systems, it does not segment lesions or pinpoint exact biopsy locations, which may limit its usefulness in guiding procedures. It also lacks adjustable diagnostic thresholds, and its knowledge may not reflect the latest data or population-specific nuances.

Ultimately, ChatGPT and traditional visual models offer complementary strengths—one provides integrative clinical reasoning, the other precise image-based analysis. Our findings suggest that even without task-specific image training, ChatGPT can perform at an expert level. This is reflected in our predictive model for CIN2+, where the clinical model (with expert judgment) achieved an AUC of 0.966, compared to 0.947 when incorporating ChatGPT (p = 0.129) (Figure 3).

A key added value of using a language model is scalability. ChatGPT, being accessible and not requiring personalized training, can be deployed in settings with few experts or limited infrastructure. It may help standardize colposcopic practice, reduce diagnostic variability, and improve screening sensitivity.

Altogether, these findings support the clinical utility of language models like ChatGPT—not only as emulators of medical expertise but as flexible platforms for decision support, training, and diagnostic equity.

This study also offers methodological strengths that enhance the reliability of its findings. First, clinical cases were systematically curated from a publicly available and recognized source: the IARC Colposcopy Atlas (18), which provides expert-interpreted images with corresponding histopathological outcomes. The study used a blinded design, with ChatGPT assessing cases independently of original diagnoses, minimizing confirmation bias. The direct comparison with pathology, when available, allowed for objective performance assessment.

Finally, this study introduces an innovative approach to AI-assisted colposcopy by leveraging a generative language model (ChatGPT-4), rather than purely image-based algorithms. Unlike traditional deep learning models trained on large datasets of cervical images, ChatGPT applies multimodal clinical reasoning by combining visual features with contextual variables like age and HPV status to generate a diagnostic decision.

Despite these strengths, some limitations must be acknowledged. The dataset used was derived from a didactic image archive, not a prospective clinical cohort, potentially limiting generalizability. Selection bias may exist, as the cases may reflect idealized or textbook patterns rather than real-world variability. Additionally, the analysis relied on static images captured after acetic acid and/or Lugol application, without the dynamic visual and tactile information provided by live colposcopy. ChatGPT assessed these asynchronously and without patient interaction. Finally, this model has not yet been prospectively validated in a clinical setting, and future studies will be needed to confirm its performance in real-world environments with direct patient care.

The findings of this study suggest that language models such as ChatGPT could serve as useful adjunctive tools in colposcopic decision-making, especially for less experienced professionals or in under-resourced settings. Its ability to synthesize clinical reasoning, interpret images, and provide explanatory conclusions in natural language makes it a promising second-opinion system that could enhance biopsy accuracy and reduce diagnostic variability.

Moreover, this approach has educational potential, offering structured clinical reasoning that could enhance colposcopy training for residents and general gynecologists. Future applications may include real-time clinical assistance and integration into computer-aided diagnostic (CAD) platforms.

Prospective, multicenter trials are warranted to validate these findings and explore the integration of additional clinical inputs such as cytology or detailed patient history. It would also be valuable to assess ChatGPT’s utility in identifying biopsy sites or predicting lesion progression over time.

## CONCLUSIONS

This study demonstrates that a generative language model like ChatGPT-4 can achieve diagnostic performance in colposcopy comparable to that of expert colposcopists, including visual impression, clinical decision-making, and histological prediction. Its high concordance with histopathological findings, along with its excellent sensitivity for high-grade lesions (CIN2+), supports its role as a potentially valuable clinical support tool.

Unlike traditional image-based systems, ChatGPT integrates clinical reasoning, enabling a more contextualized assessment of colposcopic images. Its ability to combine visual findings with clinical variables (such as age and HPV status) suggests that language models can provide added value, particularly in settings with less colposcopic expertise or limited resources.

These findings open new avenues for the integration of generative artificial intelligence into cervical cancer screening programs—both as a clinical decision support tool and as an educational resource. Although prospective studies are needed to validate its real-world application, this approach represents an innovative step toward more accurate, accessible, and standardized colposcopy.

## Author contributions

CPP conceptualized the study, curated the dataset, performed the statistical analysis, and drafted the initial manuscript. MJGRJ contributed to the study design, critically reviewed the manuscript, and provided expert input in the interpretation of results. Both authors reviewed and approved the final version of the manuscript and agree to be accountable for all aspects of the work.

## Data Availability Statement

The datasets generated and analyzed during the current study are not publicly available due to privacy and institutional policies, but are available from the corresponding author on reasonable request.

